# Rare damaging *CCR2* variants are associated with lower lifetime cardiovascular risk

**DOI:** 10.1101/2023.08.14.23294063

**Authors:** Marios K. Georgakis, Rainer Malik, Omar El Bounkari, Natalie R. Hasbani, Jiang Li, Jennifer E. Huffman, Gabrielle Shakt, Reinier W. P. Tack, Tamara N. Kimball, Yaw Asare, Alanna C. Morrison, Noah L. Tsao, Renae Judy, Braxton D. Mitchell, Huichun Xu, May E. Montasser, Ron Do, Eimear E. Kenny, Ruth J.F. Loos, James G. Terry, John Jeffrey Carr, Joshua C. Bis, Bruce M. Psaty, W. T. Longstreth, Kendra A Young, Sharon M Lutz, Michael H Cho, Jai Broome, Alyna T. Khan, Fei Fei Wang, Nancy Heard-Costa, Sudha Seshadri, Ramachandran S. Vasan, Nicholette D. Palmer, Barry I. Freedman, Donald W. Bowden, Lisa R. Yanek, Brian G. Kral, Lewis C. Becker, Patricia A. Peyser, Lawrence F. Bielak, Farah Ammous, April P. Carson, Michael E. Hall, Laura M. Raffield, Stephen S. Rich, Wendy S. Post, Russel P. Tracy, Kent D. Taylor, Xiuqing Guo, Michael C. Mahaney, Joanne E. Curran, John Blangero, Shoa L. Clarke, Jeffrey W. Haessler, Yao Hu, Themistocles L. Assimes, Charles Kooperberg, Jürgen Bernhagen, Christopher D. Anderson, Scott M. Damrauer, Ramin Zand, Jerome I. Rotter, Paul S. de Vries, Martin Dichgans

**Affiliations:** Institute for Stroke and Dementia Research (ISD), University Hospital, Ludwig-Maximilians-University (LMU), Munich, Germany; Program in Medical and Population Genetics, Broad Institute of MIT and Harvard, Cambridge, MA, USA; Munich Cluster for Systems Neurology (SyNergy), Munich, Germany; Human Genetics Center, Department of Epidemiology, Human Genetics, and Environmental Sciences, School of Public Health, The University of Texas Health Science Center at Houston, Houston, TX, USA; Department of Molecular and Functional Genomics, Geisinger Health System, Danville, Pennsylvania, USA; Veterans Affairs Healthcare System, Boston, MA, USA; Department of Surgery, Perelman School of Medicine at the University of Pennsylvania, Philadelphia, PA, USA; Corporal Michael Crescenz VA Medical Center, Philadelphia, PA, USA; Center for Genomic Medicine, Massachusetts General Hospital, Boston, MA, USA; Department of Neurology, Brigham and Women’s Hospital, Boston, MA, USA; Department of Medicine, University of Maryland School of Medicine, Baltimore, MD; Geriatrics Research and Education Clinical Center, Baltimore Veterans Administration Medical Center, Baltimore, MD; The Charles Bronfman Institute for Personalized Medicine, Icahn School of Medicine at Mount Sinai, New York, NY, USA; Department of Genetics and Genomic Sciences, Icahn School of Medicine at Mount Sinai, New York, NY, USA; The Center for Genomic Health, Icahn School of Medicine at Mount Sinai, New York, NY, USA; Pamela Sklar Division of Psychiatric Genomics, Icahn School of Medicine at Mount Sinai, New York, NY, USA; Department of Medicine, Icahn School of Medicine at Mount Sinai, New York, NY, USA; Department of Radiology, Vanderbilt University Medical Center, Nashville, TN, USA; Cardiovascular Health Research Unit, Department of Medicine, University of Washington, Seattle, WA, USA; Department of Health Systems and Population Health, University of Washington, Seattle, WA, USA; Department of Epidemiology, University of Washington, Seattle, WA, USA; Department of Neurology, University of Washington, Seattle, WA, USA; Department of Epidemiology, University of Colorado Anschutz Medical Campus, Aurora CO, USA; Department of Population Medicine, PRecisiOn Medicine Translational Research (PROMoTeR) Center, Harvard Pilgrim Health Care and Harvard Medical School, Boston, MA, USA; Department of Biostatistics, T.H. Chan School of Public Health, Harvard University, Boston, MA, USA; Channing Division of Network Medicine, Brigham and Women’s Hospital, Harvard Medical School, Boston, MA, USA; Department of Biostatistics, University of Washington, Seattle, WA, USA; Department of Medicine, Boston University School of Medicine, Boston, MA, USA; Boston University and National Heart, Lung, and Blood Institute’s Framingham Heart Study, Framingham, MA, USA; Bigg’s Institute for Alzheimer’s Disease and neurodegenerative disorders, University of Texas Health Science Center, San Antonio, TX, USA; Department of Epidemiology, Boston University School of Public Health, Boston, MA, USA; Department of Biochemistry, Wake Forest School of Medicine, Winston-Salem, NC, USA; Section on Nephrology, Department of Internal Medicine, Wake Forest School of Medicine, Winston-Salem, NC, USA; Department of Medicine, Johns Hopkins University School of Medicine, Baltimore, MD, USA; Department of Epidemiology, School of Public Health, University of Michigan, Ann Arbor, MI, USA; Department of Medicine, University of Mississippi Medical Center, Jackson, MS; Department of Genetics, University of North Carolina at Chapel Hill, Chapel Hill, NC; Center for Public Health Genomics, University of Virginia, Charlottesville, VA USA; Johns Hopkins Bloomberg School of Public Health, Johns Hopkins School of Medicine, Baltimore, MD USA; Departments of Pathology & Laboratory Medicine, and Biochemistry, Larner College of Medicine, University of Vermont, Burlington, VT USA; The Institute for Translational Genomics and Population Sciences, Department of Pediatrics, The Lundquist Institute for Biomedical Innovation at Harbor-UCLA Medical Center, Torrance, CA USA; Department of Human Genetics and South Texas Diabetes and Obesity Institute, University of Texas Rio Grande Valley School of Medicine, Brownsville TX USA; Department of Medicine (Division of Cardiovascular Medicine), Stanford University School of Medicine, Stanford, CA, USA; Stanford Cardiovascular Institute, Stanford, CA, USA; VA Palo Alto Health Care System, Palo Alto, CA, USA; Division of Public Health Sciences, Fred Hutchinson Cancer Center, Seattle WA 98109 USA; Department of Genetics, Perelman School of Medicine at the University of Pennsylvania, Philadelphia, PA, USA; Department of Neurology, Pennsylvania State University, Hershey, Pennsylvania, USA; Department of Neurology, Neuroscience Institute, Geisinger Health System, Danville, PA, USA; German Centre for Neurodegenerative Diseases (DZNE), Munich, Germany; German Centre for Cardiovascular Research (DZHK, Munich), partner site Munich Heart Alliance, Munich, Germany

**Author notes:** equally contributed (Marios K. Georgakis, Rainer Malik). **Address for Correspondence:** Marios K. Georgakis, MD, PhD, Clinician-Scientist, Martin Dichgans, MD, Director, Institute for Stroke and Dementia Research, Ludwig-Maximilians-University Hospital Feodor-Lynen-Str. 17, 81377 Munich, Germany.

**Keywords:** atherosclerosis, CCR2, genetics, inflammation, cardiovascular disease

## Abstract

**Background:** Previous work has shown a role of CCL2, a key chemokine governing monocyte trafficking, in atherosclerosis. However, it remains unknown whether targeting CCR2, the cognate receptor of CCL2, provides protection against human atherosclerotic cardiovascular disease.

**Methods:** Computationally predicted damaging or loss-of-function (REVEL>0.5) variants within *CCR2* were detected in whole-exome-sequencing data from 454,775 UK Biobank participants and tested for association with cardiovascular endpoints in gene-burden tests. Given the key role of CCR2 in monocyte mobilization, variants associated with lower monocyte count were prioritized for experimental validation. The response to CCL2 of human cells transfected with these variants was tested in migration and cAMP assays. Validated damaging variants were tested for association with cardiovascular endpoints, atherosclerosis burden, and vascular risk factors. Significant associations were replicated in six independent datasets (n=1,062,595).

**Results:** Carriers of 45 predicted damaging or loss-of-function *CCR2* variants (n=787 individuals) were at lower risk of myocardial infarction and coronary artery disease. One of these variants (M249K, n=585, 0.15% of European ancestry individuals) was associated with lower monocyte count and with both decreased downstream signaling and chemoattraction in response to CCL2. While M249K showed no association with conventional vascular risk factors, it was consistently associated with a lower risk of myocardial infarction (Odds Ratio [OR]: 0.66 95% Confidence Interval [CI]: 0.54-0.81, p=6.1×10^-5^) and coronary artery disease (OR: 0.74 95%CI: 0.63-0.87, p=2.9×10^-4^) in the UK Biobank and in six replication cohorts. In a phenome-wide association study, there was no evidence of a higher risk of infections among M249K carriers.

**Conclusions:** Carriers of an experimentally confirmed damaging *CCR2* variant are at a lower lifetime risk of myocardial infarction and coronary artery disease without carrying a higher risk of infections. Our findings provide genetic support for the translational potential of CCR2-targeting as an atheroprotective approach.

## INTRODUCTION

Atherosclerotic cardiovascular disease (CVD) is the leading cause of morbidity and mortality worldwide.^1–3^ Over 20 years of preclinical research have provided overwhelming evidence for a causal role of inflammation in atherogenesis^4,5^ and recent trials provided proof-of-concept that targeting inflammation can lead to reductions in adverse cardiovascular events.^6–8^ The canakinumab anti-inflammatory thrombosis outcome study (CANTOS) demonstrated that treatment with a monoclonal antibody against IL-1β lowers risk of recurrent vascular events among individuals with recent myocardial infarction.^6^ The colchicine cardiovascular outcomes trial (COLCOT)^7^ and the low-dose colchicine-2 (LoDoCo2) trial^8^ further showed that colchicine, an established drug with widespread inhibitory effects on inflammatory pathways,^9,10^ lowers the risk of recurrent vascular events in patients with coronary artery disease (CAD). Targeting inflammation for atheroprotection must be balanced against the impact on any host defense responses. For example, both canakinumab^6^ and colchicine^7^ were associated with adverse effects including fatal infections in the CANTOS and the colchicine trials. While translational efforts have mostly focused on the inflammasome-IL-1β/IL-6 axis,^11^ evidence from preclinical studies and early-phase clinical trials highlights the promise of alternative cytokines^5^ for the development of a second generation of atherosclerosis-centered anti-inflammatory treatments.^4^

CC-motif chemokine ligand 2 (CCL2) is a pivotal inflammatory chemokine regulating monocyte trafficking^12^ that has been studied as a potential target in atherosclerosis. Preclinical data suggest that pharmacological targeting of CCL2 or its receptor CCR2 might lower atherosclerosis burden in experimental models.^13^ However, only recently large-scale genetic and epidemiological studies have highlighted the relevance of the CCL2/CCR2 pathway in human CVD, calling for clinical translation of strategies targeting this pathway.^14^ Prospective observational studies^15,16^ support associations of higher circulating CCL2 levels with ischemic stroke and cardiovascular death, whereas Mendelian randomization analyses from population genetic studies^17,18^ show associations with higher risk of ischemic stroke and coronary artery disease. Furthermore, CCL2 levels are higher in atherosclerotic lesions from patients with symptomatic carotid stenosis, as compared to asymptomatic disease and are associated with features of plaque vulnerability.^19^

Although these studies support a role of the CCL2/CCR2 axis in human atherosclerosis, it remains unclear whether pharmacological intervention in this pathway could lead to atheroprotection in humans. Several molecules targeting CCR2 are currently under development for autoimmune disease, liver disease, and cancer and could be repurposed for prevention of atherosclerotic cardiovascular disease.^14^ Studies examining the phenotypic effects of rare genetic variants in population-based studies have been instrumental in predicting the consequences of pharmacological interventions^20–24^ and might thus serve as a validation step for drug targets under development.

Here, we leveraged whole-exome sequencing (WES) data from 454,775 participants of the population-based UK Biobank (UKB) study to explore whether rare damaging variants in the *CCR2* gene are associated with cardiovascular disease risk, burden of atherosclerosis, and traditional vascular risk biomarkers. We experimentally confirmed the damaging effects of one variant (M249K), which has a frequency of 0.15% among UKB participants of European ancestry and replicated its effects on cardiovascular risk in external population- and hospital-based biobanks. Finally, we performed a phenome-wide association study across two biobanks to explore associations of this damaging variant with the risk of infections or other potential safety signals of any CCR2-targeting treatments.

## METHODS

### Study population

We used data from the UKB, a population-based prospective cohort study of UK residents aged 40-69 years recruited between 2006-2010 from 22 assessment centers across the UK.^25^ This analysis was restricted to 454,775 out of 502,419 participants with available whole-exome sequencing data (UKB Exome 450k release from October 2021). Primary and secondary analyses were performed with an updated Functional Equivalence (FE) protocol that retains original quality scores in the CRAM files (referred to as the OQFE protocol). We included only variants that met published criteria^26^: individual and variant missingness <10%, Hardy Weinberg Equilibrium p-value>10^-15^, minimum read coverage depth of 7 for SNPs and 10 for indels, at least one sample per site passed the allele balance threshold >0.15 for SNPs and 0.20 for indels. We used genotype array data released by the UKB study to assign individuals to continental ancestry super-groups (African (AFR), Hispanic or Latin American (HLA, originally referred to as ‘AMR’ by the 1000 Genomes Project), East Asian (EAS), European (EUR) and South Asian (SAS)) by projecting each sample onto reference principal components (PCs) calculated from the 1000 Genomes reference panel. We included only individuals without evidence of relatedness within the UKB samples, as defined by a KING cut-off of <0.084.

### Detection of damaging genetic variants in *CCR2* and experimental validation

Restricting our analyses to the *CCR2* gene, we detected predicted LoF or damaging missense variants with a MAF<1% in the *CCR2* exonic region and then explored which of those are associated with monocyte count as a functional readout. Variants from WES were annotated as previously described^27^ using VEP v101.^28,29^ We used the LofTee plugin for predicting LoF variants^30^ and a REVEL cutoff of >0.5 from dbNSFP version 4.0a for predicted damaging missense mutations.

We aimed to further restrict our analyses to damaging variants with proven functional effects. One of the key functions of the CCL2 axis is the recruitment of classical monocytes from the bone marrow to the circulation in a CCR2-dependent way.^31–33^ Consequently, damaging variants in the *CCR2* gene would be expected to be associated with a lower monocyte count in the circulation. To test associations with the circulating counts of monocytes and other white blood cells (WBC), absolute counts were extracted from the UKB fields *30160* (basophil counts), *30150* (eosinophil counts), *30130* (monocyte counts), *30120* (lymphocyte counts) and *30140* (neutrophil counts). Distributions were visually checked for normal distribution and log-normalized when needed. Associations were tested using regenie v2.2.4.^34^ For WBC analyses, we used sex, age at blood draw, and the first 5 ancestral PCs as covariates. The mixed model parameters were estimated using 200,000 genotyped common variants. Saddle point approximation regression was applied. Genetic variants associated with monocyte count at a false discovery rate (FDR)-corrected p-value <0.05 were prioritized for further experimental validation. To test whether the associations are specific for monocyte count, we also tested associations with other WBC counts (basophils, eosinophils, lymphocytes, neutrophils) for each variant.

Two predicted damaging CCR2 variants showing a significant association with lower monocyte count were brought forward to experimental testing. The two variants were M249K (3:46358273:T:A) and W165S (3:46358021:G:C, both hg38). First, we tested how human *CCR2*-knockout monocytic THP-1 cells transfected with these variants or wild-type *CCR2* respond to chemoattraction by CCL2 using a Transwell migration assay (details in **Supplementary Methods**). Next, we tested whether M249K-mutant *CCR2* (which showed an effect in the migration assay) influences cAMP production in response to CCL2 in transfected human HEK293T cells (details in **Supplementary Methods**).

### Associations with cardiovascular risk, atherosclerosis burden and vascular risk factors

Focusing on two sets of variants, we then explored associations of CCR2 variants with clinical cardiovascular endpoints including myocardial infarction, coronary artery disease, acute ischemic stroke, peripheral artery disease, and abdominal aortic aneurysm using ICD-10- and ICD-9-coded diagnoses, OPCS4-coded procedures, self-report, and algorithmically-defined phenotypes provided by the UKB as detailed in **Supplementary Table S1**. As our hypothesis was that these variants would lead to reductions in risk by lowering the lifetime burden of atherosclerosis, we also constructed a combined phenotype of atherosclerosis manifestations in four vascular beds (coronary arteries, cerebrovascular system, peripheral arteries of the extremities, and aortic atherosclerosis). Severe atherosclerosis was defined as presence of clinical manifestations in at least two vascular beds (**Supplementary Table S1**).

We followed two approaches to test associations with these phenotypes: (i) we performed a burden test combining all predicted damaging variants (LoF or REVEL >0.5) within *CCR2*; (ii) focusing on M249K, the only damaging monocyte-lowering *CCR2* variant that was experimentally validated in the migration and cAMP assays, we performed logistic regression analysis adjusted for age, sex, and the first 5 ancestral PCs testing. We included both prevalent and incident endpoints as outcomes in all analyses and used Firth‘s correction for unbalanced case/control ratios in our logistic regression analysis. To explore whether the associations with cardiovascular risk are mediated through known vascular risk factors, we further tested associations of the damaging genetic variants with the following phenotypes: systolic and diastolic blood pressure, circulating LDL and HDL cholesterol, circulating apolipoprotein B levels, circulating glycated hemoglobin A1c (HbA1c) concentration, body mass index, and C-reactive protein. Individuals under antihypertensive medications were excluded in the analyses for systolic and diastolic blood pressure, individuals under lipid-lowering medications were excluded from the analyses for LDL and HDL cholesterol and apolipoprotein B levels, and individuals under glucose-lowering treatments were excluded from the analyses for HbA1c.

### Validation of M249K *CCR2* variant in external datasets and meta-analysis

For external validation, we obtained summary statistics from six different data sources based on a pre-defined protocol: the TransOmics and Precision Medicine Program supported by NHLBI (TOPMed) Program including multiple studies and a trans-ancestry population in the US (n=51,732), the population-based deCODE dataset in Iceland (n=345,992), the hospital-based Penn Medicine Biobank (PMBB, n=43,721), the Million Veteran Programme (MVP, n=438,905), the Geisinger DiscovEHR-MyCode cohort in central and northeastern Pennsylvania (n=145,826), as well as the hospital-based Mass General Brigham Biobank (MGBB, n=36,419).

Specifically, we requested data from logistic regression models adjusted for age, sex, race, study-specific variables (e.g. sequencing center) and the first 10 PCs (with the Firth’s correction) for the M249K variant. The variant was either assessed by sequencing or directly genotyped in all cohorts. Details about the individual cohorts are provided in **Supplementary Methods**. The derived odds ratios (OR) from the six datasets were meta-analyzed using fixed-effects and random-effects meta-analyses and were subsequently also meta-analyzed with the results from the UKB. We set a significance threshold of p<0.05 in the replication meta-analysis. Heterogeneity was assessed with the I^2^ and the Cochran Q statistic.

### Phenome-wide association study

To explore potential adverse effects associated with damaging *CCR2* genetic variants, we tested associations of M249K with the full range of clinical phenotypes encoded in the UKB and replicated in an external cohort (Geisinger DiscovEHR-MyCode). We used the Phecode Map 1.2 to map UKB ICD10-codes to Phecodes^35^ using all ICD10 codes (main position, secondary position, death records) from the UKB. We excluded Phecodes with <100 cases and Phecodes that are male- or female-specific. Individuals were assigned a case status if >1 ICD10 code was mapped to the respective Phecode. To approximate effect size in a logistic regression framework, we used minor allele carrier status as an independent variable and age at baseline, sex and 5 ancestry PCs as covariates. We used Firth‘s correction for unbalanced case/control ratios in our logistic regression analysis for all results with p<0.05. The results from the two cohorts were meta-analyzed using fixed- and random-effects models. Heterogeneity was assessed with the I^2^ and the Cochran Q statistic.

### Ethics and data availability

All studies have received ethical approvals from the respective ethical authorities and participants in all studies have provided written informed consent. Data from the UKB are available upon submission and approval of a research proposal. The UKB has institutional review board approval from the Northwest Multi-Center Research Ethics Committee (Manchester, UK). We accessed the data following approval of an application by the UKB Ethics and Governance Council (Application No. 36993 and 2532). Summary results from the external cohorts were obtained following a common pre-defined research protocol to principal investigators of the study and are presented in the figures and supplementary tables. deCODE has been approved by the National Bioethics Committee of Iceland, and the study was conducted in agreement with conditions issued by the Data Protection Authority of Iceland (VSN_14-015). All common research protocols for the TOPMed Program have been approved by the institutional review board at the University of Maryland Baltimore, whereas individual participating studies have obtained ethical approval from their local ethical authorities, as described previously.^36^ The Penn Medicine BioBank is approved by the University of Pennsylvania, MVP by the Veterans Affairs Central Institutional Review Board, the Geisinger DiscovEHR-MyCode cohort by Geisinger Institutional Review Boards, and MGBB by the Ethics Committee of Mass General Brigham.

## RESULTS

### Computationally predicted damaging *CCR2* variants

Among 428,191 unrelated (out of 454,775 with whole-exome sequencing data) UK Biobank participants from the whole-exome sequencing data release, we found a total of 45 predicted LoF or damaging (REVEL>0.5) variants in the exonic region of the *CCR2* gene distributed across 787 heterozygous carriers (frequency 0.18%, **Figure 1** and **Supplementary Table S2**). There was no homozygous carrier of any of these variants and variants were predominantly prevalent in individuals of European ancestry (779 carriers, frequency 0.20%), as compared to individuals of African, Hispanic or Latin American, East Asian, and South Asian ancestry (total of 8 carriers, pooled frequency 0.02%). Due to the very low frequency in other ancestries, we restricted our analyses to individuals of European ancestry (n=393,416), Baseline characteristics of the study population are presented in **Supplementary Table S3**.

**Figure 1.**
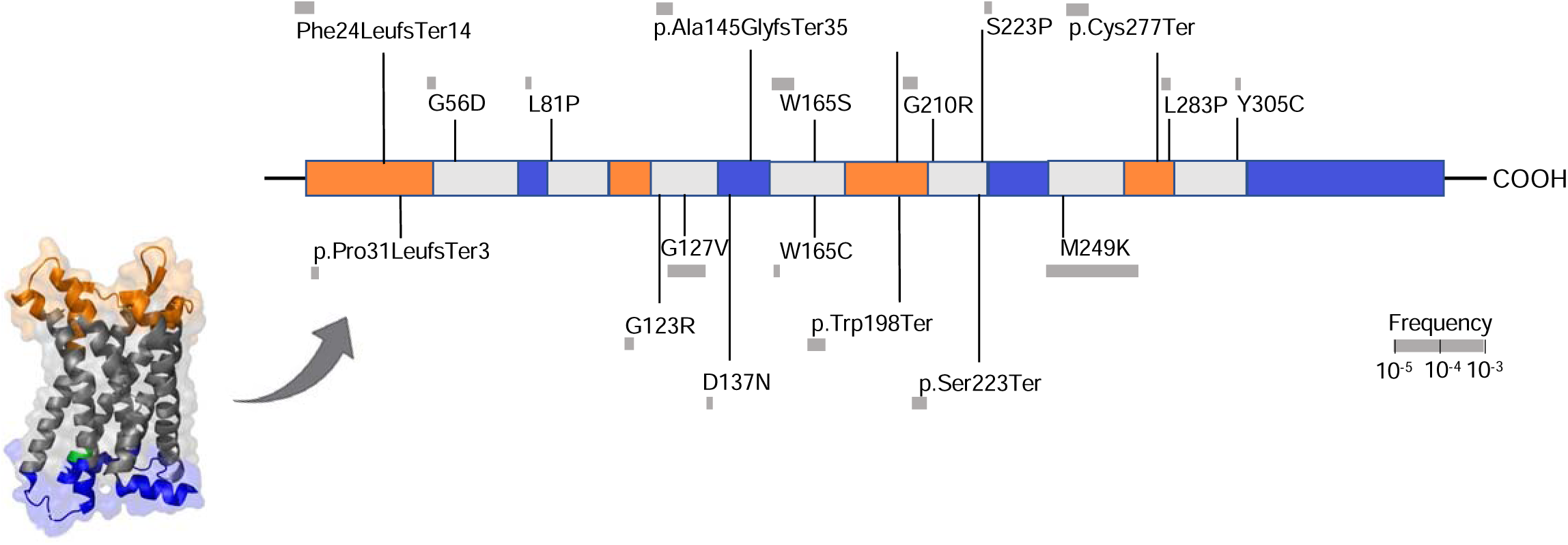
Damaging *CCR2* variants. Domain structure of the CCR2 protein and position of the predicted loss-of-function (LoF) or missense damaging (REVEL>0.5) variants present in >2 UK Biobank participants in the *CCR2* exonic regions.

### Associations of *CCR2* variants with cardiovascular endpoints in gene-burden tests

In a discovery analysis including all 45 computationally predicted LoF or damaging variants, we next explored whether genetic variation in *CCR2* is associated with atherosclerotic disease using a gene-based burden test. We found nominally significant associations of the predicted damaging *CCR2* variants with myocardial infarction (OR: 0.60, 95%CI: 0.40-0.90, p=0.008) and coronary artery disease (OR: 0.76, 95%CI: 0.59-0.99, p=0.03), as well as directionally consistent associations with the odds of all other examined outcomes (ischemic stroke, peripheral artery disease, abdominal aortic aneurysm, **Figure 2A**). Variant burden was associated with lower risk of a combined atherosclerotic endpoint (OR: 0.80, 95%CI: 0.65-0.98, p=0.03), as well as with severe atherosclerotic disease, defined by clinical manifestations in at least two vascular beds (OR: 0.24, 95%CI: 0.07-0.82 p=0.003; **Figure 2B**). We found a trend for a dose-response pattern with lower frequency of clinically manifest atherosclerotic disease across the number of vascular beds involved among carriers for these *CCR2* variants (frequency of atherosclerosis presence among carriers vs. non-carriers: in no vascular bed 88.0% vs. 90.7%; in 1 vascular bed 10.5% vs. 9.1%; in 2 vascular beds 1.2% vs. 0.3%; in 3 vascular beds 0.3% vs 0.1%, in 4 vascular beds 0.03% vs 0%; OR from ordinal regression: 0.74, 95%CI: 0.54-0.97, p=0.01).

**Figure 2.**
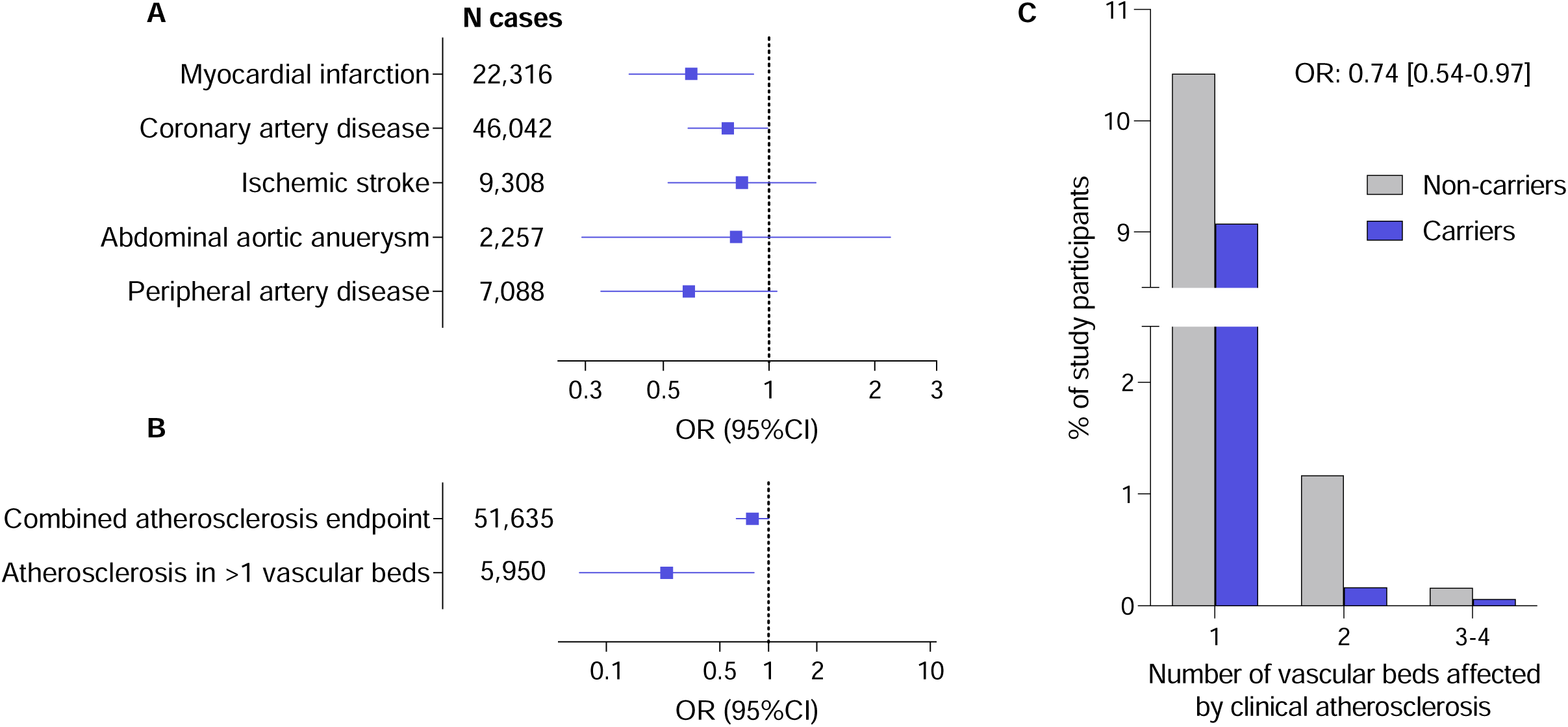
Associations of rare computationally predicted damaging *CCR2* variants with risk of atherosclerotic cardiovascular disease in the UK Biobank. Associations of 45 predicted loss-of-function (LoF) or damaging (REVEL>0.5) *CCR2* variants (minimum allele frequency <0.01) in burden tests with (**A**) risk of cardiovascular disease endpoints and (**B**) a combined atherosclerosis endpoint as well as severe clinical atherosclerotic disease defined by manifestations in at least 2 vascular beds (coronary arteries, cerebrovascular system, peripheral arteries of extremities, aorta) among UK Biobank participants of European ancestry. (**C**) Prevalence of clinically manifest atherosclerosis across 4 vascular beds (coronary arteries, cerebrovascular system, peripheral arteries of extremities, aorta) among carriers and non-carriers of the 45 predicted LoF or damaging CCR2 variants. The odds ratio (OR) is derived from ordinal regression adjusted for age, sex, and the first 5 ancestral principal components.

### Functional consequences of *CCR2* damaging variants

To move beyond computationally predicted functional effects, we followed a two-step approach to select *CCR2* variants with functional relevance. Since one of the key functions of CCL2 is the CCR2-dependent recruitment of classical monocytes from the bone marrow to the circulation, we first tested the association of the predicted damaging variants in *CCR2* with circulating monocyte counts. We found two variants (M249K and W165S) to be associated with lower monocyte counts FDR-corrected p<0.05, **Supplementary Table S2**) and prioritized them for experimental validation.

We tested whether transfection of human *CCR2*-knockout THP-1 monocytes with either of these two variants influences directed migration towards CCL2 in a trans-migration cell assay. Cells transfected with M249K showed a clearly reduced response to CCL2, when compared to cells transfected with wild-type *CCR2*, resembling the response of *CCR2*-knockout cells (**Figure 3A** and **Supplementary Figure S1)**. Cells transfected with W165S showed a profile similar to cells transfected with wild-type *CCR2* (**Supplementary Figure S1)**. HEK293T cells transfected with M249K further showed a profound reduction of cAMP production in response to CCL2, when compared to HEK293T cells transfected with wild-type *CCR2* (**Figure 3B**).

**Figure 3.**
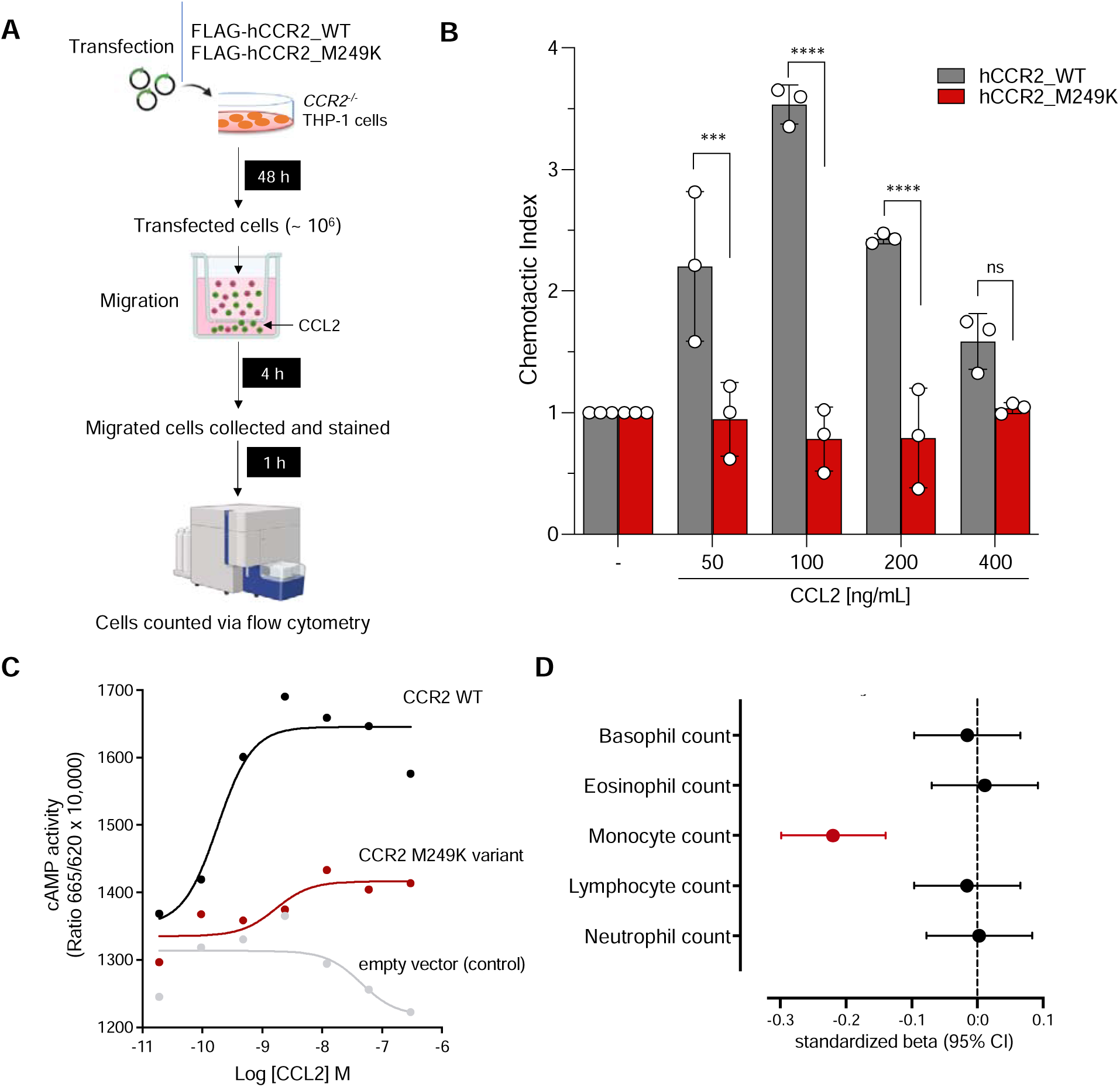
Validation of the damaging effect of M249K (3:46358273:T:A) on CCL2-driven monocyte chemotaxis. (**A**) Experimental outline of chemotaxis assay. Transfected CCR2–/– THP-1 cells (marked in green) and non-transfected cells (marked in red) were used in a Transwell chemotaxis assay. The cells that moved to the lower chamber were collected, stained, and quantified using flow cytometry. (**B**) Chemotaxis in response to increasing concentrations of CCL2 for human *CCR2*-knockout monocytic THP-1 cells transfected with the mutant M249K vs. wild-type CCR2, as determined in a trans-well migration assay (comparisons derived from two-way ANOVA, ns: p>0.05, *** p<0.001, ****p<0.0001). (**C**) Results of cyclic AMP (cAMP) assay. Shown is the cAMP activity in HEK293T cells transfected with either empty vector or wild-type or M249K *CCR2* in response to different concentrations of CCL2. Results are presented as “Ratio 665/620 x 10,000” (ratio of fluorescence at 665 nm and 620 nm x 10,000). (**D**) Associations of M249K with counts of different leukocyte populations in the subset of European ancestry participants of the UK Biobank (N=393,838), as derived from linear regression models adjusted for age, sex, and the first 5 ancestral principal components.

M249K has a frequency of 0.15% among UKB participants of European ancestry and leads to the replacement of methionine by lysine in the sixth transmembrane domain of the CCR2 receptor (**Figure 1**). Confirming the specificity of the consequences of M249K on monocyte recruitment, we found no evidence of associations with any of the other leukocyte type counts in the UKB (**Figure 3C**).

### Associations of M249K with cardiovascular risk and replication in external cohorts

Carriers of the M249K variant were at lower risk of myocardial infarction (OR: 0.62 95%CI: 0.40-0.97, p=0.03) and coronary artery disease (OR: 0.73 95%CI: 0.56-0.97, p=0.02) in the UKB (**Figure 4A)**. As expected, given that M249K was by far the most frequent among computationally predicted damaging CCR2 variants, we found associations with a lower risk of the combined atherosclerosis endpoint, as well as significantly lower risk of severe atherosclerosis (manifestations in ≥2 vascular beds, **Figure 4B**), similar to the burden test. To explore whether the effects of M249K are mediated through effects on risk factors targeted by current preventive approaches, we next tested associations with established biomarkers of vascular risk. We found no associations with any of the tested vascular risk factors including blood pressure, hyperglycemia, or circulating lipids. Furthermore, there was no evidence of association with the levels of the known inflammatory biomarker C-reactive protein, thus suggesting that the effects might be independent of factors targeted by current atheroprotective anti-inflammatory treatments (**Figure 4C**).

**Figure 4.**
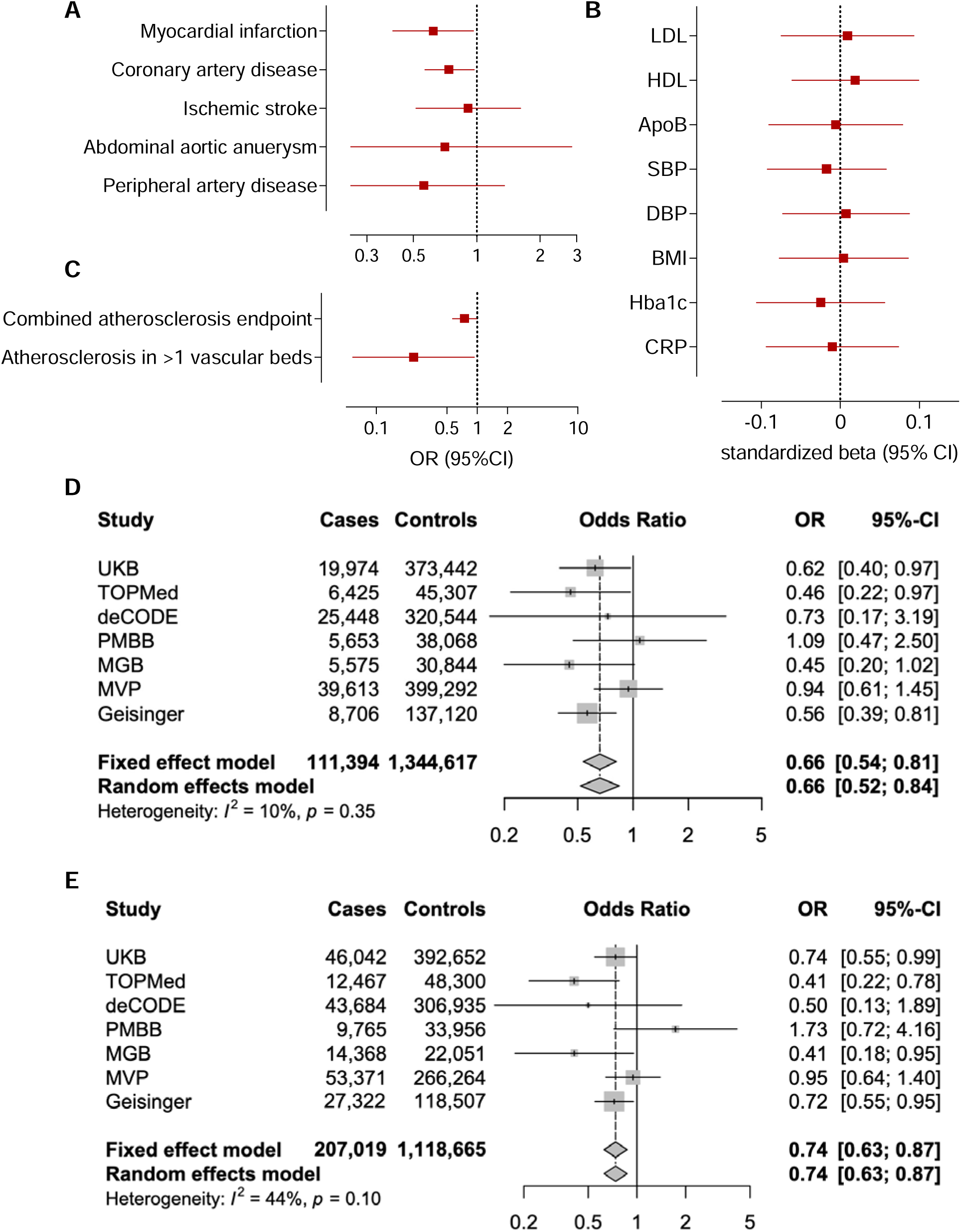
Association of the damaging M249K *CCR2* variant with lifetime cardiovascular risk. Associations of the M249K *CCR2* variant with risk of **(A)** cardiovascular disease endpoints, (**B**) a combined atherosclerosis endpoint and severe clinical atherosclerotic disease defined by manifestations in at least 2 vascular beds (coronary arteries, cerebrovascular system, peripheral arteries of extremities, aorta), and (**C**) conventional vascular risk biomarkers in models adjusted for age, sex, and the first five principal components among UK Biobank participants of European ancestry. HbA1c and C-reactive protein levels are log-transformed for normalization. Fixed-effects and random-effects meta-analysis of the effects of M249K on risk of (**D**) myocardial infarction and (**E**) coronary artery disease across 7 population- and hospital-based biobanks. The effects correspond to odds ratios (OR) derived from logistic regression models adjusted for age, sex, and the first 5 ancestral principal components in each biobank.

**Figure 5.**
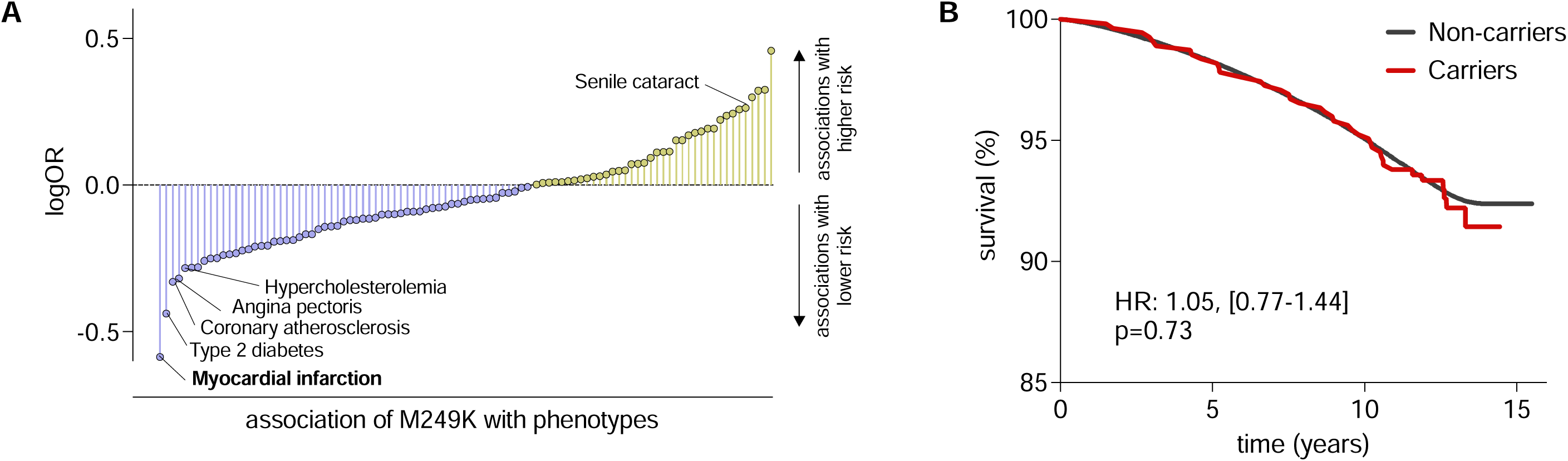
Phenome-wide association study of the damaging M249K *CCR2* variant and associations with overall survival. (**A**) Results from a phenome-wide association study in UK Biobank and the Geisinger DiscovEHR-MyCode cohort for the M249K *CCR2* variant amongparticipants of European ancestry. Only phecodes with ≥10 cases in UK Biobank were analyzed. We present the names of phenotypes associated with rare CCR2 variants at p<0.05. Only myocardial infarction reached an FDR-corrected p-value of <0.05. The results are presented as log-Odds Ratios (log-OR). (**B**) Kaplan-Meier curves for overall survival across 15 years of follow-up among carriers and non-carriers of the M249K *CCR2* variant in the European subset of UK Biobank participants.

As these associations were only nominally significant and suggestive of an effect of this variant, we aimed to externally replicate our findings to provide robust evidence for an association with a lower risk of myocardial infarction and coronary artery disease. To externally replicate the effects of M249K, we meta-analyzed data from six external datasets (TOPMed, deCODE, PMBB, MGB, MVP, Geisinger). The frequency of the variant varied between the six datasets (range: 0.0006% in MVP to 0.228% in Geisinger), totaling 767 heterozygous carriers among 1,062,595 individuals (91,420 myocardial infarction and 142,361 coronary artery disease cases). When meta-analyzing data from the six cohorts for the association between carrying M249K and risk of myocardial infarction or coronary artery disease, the ORs were comparable to those of the UKB (OR for myocardial infarction: 0.67 95%CI: 0.53-0.85, p=0.0006; OR for coronary artery disease: 0.74 95%CI: 0.53-0.85, p=0.003, **Supplementary Figure S2** and **Supplementary Table S4**). When meta-analyzing these data with UKB reaching a sample size of 1,456,011 individuals including 1,314 M249K carriers (111,394 and 207,019 cases of myocardial infarction and coronary artery disease, respectively), we found M249K carriers to have 34% lower odds of suffering a myocardial infarction (OR: 0.66 95%CI: 0.54-0.81, p=6.1×10^-5^) and 26% lower odds of coronary artery disease (OR: 0.74 95%CI: 0.63-0.87, p=2.9×10^-4^, **Figure 4D-E**, **Supplementary Table S4**).

### Phenome-wide association study

As a last step, we performed a PheWAS to explore whether M249K is associated with potential adverse effects that would raise signals for possible side-effects of any CCR2-targeting treatments. Because we lacked statistical power for most of the outcomes, we restricted our analyses to endpoints with ≥10 cases in the carrier group of the UKB. To increase statistical power, we meta-analyzed our results with the Geisinger cohort. Across all phenotypes only myocardial infarction showed an association with M249K that passed the FDR-corrected p<0.05 threshold. Coronary atherosclerosis and angina pectoris were also less frequent (p<0.05) among carriers although only at a nominal non-corrected p<0.05 (**Figure 4A**). No infectious disease phenotypes were found to be more common (p<0.05) among M249K carriers (**Supplementary Table S5**). Senile cataract was more common among carriers (p<0.05). Among UKB participants, we found no evidence for an association between carrier status for a *CCR2* damaging variants and survival over a 15-year follow-up period (**Figure 4B**).

## DISCUSSION

Using data from 1.5M individuals we found that heterozygous carriers of a rare experimentally confirmed damaging *CCR2* variant (M249K) are at lower lifetime risk of myocardial infarction and coronary artery disease. Carriers of this variant showed no differences in LDL cholesterol, blood pressure, BMI, HbA1c and C-reactive protein levels, indicating that damaging *CCR2* variants exert their effects independently of conventional vascular risk factors targeted by available atheroprotective treatments. On the other hand, we found no evidence of associations with higher risk of infectious diseases or overall mortality among carriers. Collectively, our results provide genetic support for CCR2 as a potential therapeutic target for atherosclerotic cardiovascular disease.

Our findings extend our previous results from a Mendelian randomization study suggesting a higher atherosclerosis risk among individuals with genetic predisposition to elevated CCL2 levels.^17^ That study focused on common genetic variants distributed across the genome and shown to influence CCL2 levels in *trans*. In contrast, the current study assessed damaging rare genetic variants within the *CCR2* gene. The functional impact of M249K, the most frequent variant among those associated with atherosclerotic disease, was experimentally confirmed. As such, our results are unlikely to be confounded by pleiotropic effects of the variants. Despite the lack of data from clinical trials targeting CCR2 in patients with cardiovascular disease, studies in experimental models of atherosclerosis have provided supportive evidence for an atheroprotective effect of pharmacologically targeting CCR2.^13^

Our work consistently connects these findings to humans and provides a rationale for conducting a clinical trial with pharmacological targeting of CCR2 in patients with atherosclerosis. Ultimately, only clinical testing can provide evidence in favor of or against pharmacological targeting of CCR2 for patients with atherosclerosis. A major bottleneck for initiating a clinical trial would be the need for proof of concept in a phase 2 trial before a trial with hard endpoint is conducted. Given the evidence that CRP levels are not directly affected by the CCL2/CCR2 axis,^15^ an intermediate endpoint is needed that captures plaque inflammation, and in particular, macrophage accumulation.^13^ PET imaging, particularly with an FDG tracer, has shown promise for detecting macrophage content in large plaques and has already been used in phase 2 trials targeting plaque inflammation.^37–39^ In addition, recent advances in tracer development have enabled the development of a tracer that specifically binds to CCR2.^40^ Initial data from patients have shown promise in imaging monocyte recruitment to the heart following myocardial infarction.^40^

Interestingly, we found no evidence supporting a connection of *M249K* with established pharmacological targets of atherosclerosis, such as LDL cholesterol, hyperglycemia, or elevated blood pressure. This implies that any benefit from CCR2-targeting approaches would be expected to be independent of available preventive approaches against atherosclerotic disease. Also, we found no evidence of associations with circulating C-reactive protein levels, a biomarker of the activity of the IL-6 signaling pathway that is also the target of atheroprotective anti-inflammatory treatments currently under development.^41^ Based on this, our findings suggest that CCR2 inhibition might have an atheroprotective effect on top of such approaches, which in turn might be an argument to pursue approaches embracing combination therapies.

Our PheWAS analysis was limited by low statistical power. Nevertheless, it is noteworthy that we found no evidence for an association of M249K with risk of infections, which is a potential barrier to the use of atheroprotective anti-inflammatory treatments. This is in line with previous early phase clinical trials testing inhibitors of CCR2 for other indications, which also found no important safety concerns.^42–44^ Furthermore, we found no evidence of associations with overall survival, minimizing concerns about a significant impact on unknown fatal adverse effects. The reason why we do not see a protective effect on survival despite the effect on risk of myocardial infarction could be related to statistical power and length of follow-up. It is a common observation that medications with strong effects on lowering cardiovascular risk, such as statins, have failed to show survival benefits.

Our study has limitations. First, despite leveraging the largest available whole-exome sequencing studies totaling almost 1.5 million individuals, our results for the PheWAS analyses are still limited by low statistical power and should be interpreted cautiously. Given the rarity of the damaging *CCR2* variants, any association with rare potential side-effects would ultimately be undetectable in the context of this study. While formal statistical replication is precluded by this power issue, the consistent effect directions and the overall significance of the meta-analysis raise confidence that the identified association is valid. Further, our main results for associations between M249K and myocardial infarction or coronary artery disease failed to reach the formal thresholds of statistical significance for gene-based or variant-based testing (2.7×10^-6^ and 5×10^-8^, respectively). As such, this result should be interpreted as suggestive. Genetic analyses with rare variants are still underpowered, as also highlighted in recent burden test studies from the UK Biobank, where no gene showed significant associations with either myocardial infarction or ischemic stroke.^45^ Future research should aim to pool additional data as they become available. Second, UK Biobank consists primarily of European individuals, and consequently, we detected damaging *CCR2* variants that are predominantly detected in European populations. As such, our results should not be extended to non-European individuals. Third, all analyses are based on individuals heterozygous for the damaging *CCR2* variants, as we found no homozygotes for damaging *CCR2* variants. While this lack of homozygotes can be fully explained by the frequency of damaging *CCR2* variants in the general population, it remains unknown whether homozygous status for damaging CCR2 variants would lead to potentially fatal complications. Fourth, the data from the UKB do not allow a distinction to be made between classical and non-classical monocytes. The available data suggest that CCL2 only acts on classical CCR2+ monocytes in humans or Ly6C^hi^ monocytes in mice. Experimental studies in mice have revealed that *Ccr2*-deficient Ly6C^hi^ monocytes are trapped in the bone marrow and are not mobilized after infections.^31^ Future studies in humans should further explore the impact of the examined mutations on monocytes subtypes. However, it should be noted that inhibition of monocyte recruitment from the circulation into atherosclerotic lesions is the main suspected mechanism by which CCR2 damaging variants act, and the effects of lowering monocyte counts remain to be tested.

In conclusion, we found that heterozygous carriers of a rare damaging *CCR2* variant are at lower lifetime risk of myocardial infarction and coronary artery disease without carrying a higher risk of infections. This provides further genetic support for the concept that pharmacological targeting of CCR2 might be an efficient and viable immunotherapeutic strategy to prevent atherosclerotic cardiovascular disease.

## Data Availability

Data from the UKB are available upon submission and approval of a research proposal.

## Acknowledgements

Marios K. Georgakis is supported with a clinician-scientist grant from the Munich Cluster for Systems Neurology (EXC 2145 SyNergy, ID 390857198), and an Emmy Noether grant (GZ: GE 3461/2-1, ID 512461526) by the German Research Foundation (Deutsche Forschungsgemeinschaft, DFG), as well as by the Fritz-Thyssen Foundation (Ref. 10.22.2.024MN) and the Hertie Foundation (Hertie Network of Excellence in Clinical Neuroscience, ID P1230035). Scott M. Damrauer is supported by IK2-CX001780. We acknowledge the Penn Medicine BioBank (PMBB) for providing data and thank the patient-participants of Penn Medicine who consented to participate in this research program. We would also like to thank the Penn Medicine BioBank team and Regeneron Genetics Center for providing genetic variant data for analysis. The PMBB is approved under IRB protocol# 813913 and supported by Perelman School of Medicine at University of Pennsylvania, a gift from the Smilow family, and the National Center for Advancing Translational Sciences of the National Institutes of Health under CTSA award number UL1TR001878. We thank Geisinger MyCode Community Health Initiative and Geisinger-Regeneron DiscovEHR collaboration contributors who have been critical in the generation of the original data used for this study. This publication does not represent the views of the Department of Veterans Affairs or the United States Government. Paul S. de Vries, Natalie R. Hasbani, and Alanna C. Morrison were supported by R01 HL146860. Joshua C. Bis and Bruce M. Psaty were funded in part by R01HL105756. Reading of the carotid IMT measures was supported by N01HC85085, N01HC45133; Coronary Calcium scans were supported by R01HL64587. Kendra A. Young is supported by NHLBIn U01 HL089897 and U01 HL089856. Sharon M. Lutz was supported by R01 MH129337. Nicholette D. Palmer, Barry I. Freedman and Donald W. Bowden were supported by R01 AG058921. Patricia A. Peyser was supported by R01 HL146860. Lawence F. Bielak was supported by R01 HL146860. LMR was supported by T32 HL129982 and by the National Center for Advancing Translational Sciences, National Institutes of Health, through Grant KL2TR002490. Christopher D. Anderson receives sponsored research support from the US National Institutes of Health (R01NS103924, U01NS069673), the American Heart Association (AHA 21SFRN812095). Whole genome sequencing (WGS) for the Trans-Omics in Precision Medicine (TOPMed) program was supported by the National Heart, Lung and Blood Institute (NHLBI). Cohort-specific acknowledgements for studies included in the TOPMed program are provided in **Supplementary Table S6**. Jürgen Bernhagen is supported by grants from the Deutsche Forschungsgemeinschaft (DFG; CRC 1123 [A3], BE 1977/14-1, and Munich Cluster for Systems Neurology [SyNergy, EXC 2145; ID390857198]. Martin Dichgans is supported by grants from the Deutsche Forschungsgemeinschaft (DFG; CRC 1123 [B3], DI-722/16-1 [ID: 428668490], DI-722/21-1, and Munich Cluster for Systems Neurology [SyNergy, EXC 2145; ID390857198]), the Leducq Foundation, the flagship P4-medicine project *DigiMe*d Bayern and the Vascular Dementia Research Foundation.

