## Supplementary Methods for "Rare damaging *CCR2* variants are associated with lower lifetime cardiovascular risk"

**Experimental validation of monocyte-lowering predicted damaging *CCR2* variants**

*Chemotaxis assay*

The cloning of N-terminal FLAG-tagged CCR2 (FLAG-hCCR2_WT) and its variants FLAG-hCCR2_M249K and FLAG-hCCR2_W165S was conducted by Eurofins Genomics (Munich, Germany). In brief, the cDNA sequences encoding human CCR2 (WT) and its variants (hCCR2_M249K, hCCR2_W165S) were synthesized. Following synthesis, these genes were inserted into *BamH*I/*Not*I-double-digested N-terminal FLAG-tagged pcDNA3.1 plasmid, generously provided by Bernard Lüscher (RWTH Aachen University, Germany). Subsequently, the generated clones were validated through Sanger sequencing.

Human wild type (Abcam, Ltd., #ab275477) and *CCR2* knockout (Abcam, Ltd., #ab273716) THP-1 monocytes were cultured in RPMI 1640 medium supplemented with 10% FBS, 0.05 mM beta-mercaptoethanol and 1% and 1% penicillin-streptomycin (P/S). *CCR2* knockout THP-1 cells were transfected with 2.5 µg FLAG-hCCR2_WT or its variants hCCR2_M249K and hCCR2_W165S) using Lipofectamine^TM^ LTX reagent (company, #15338100) following the manufacturer’s protocol. Low-passage cells with greater than 90% viability was used for the transfections. To enhance transfection efficiency and minimize cytotoxicity, transfections were performed in the presence of PLUS^TM^ Reagent and in a medium without antibiotics. Cells were then used for transmigration assays 18-20 hours after transfection.

The chemotactic migration of human wild type (#ab275477) and transfected or untransfected *CCR*2 knockout (#ab273716) THP-1 monocytes was assessed using a Transwell-based assay as described previously ^1^. Initially, cells were cultured in RPMI 1640 medium supplemented with 2% FBS and 1% P/S at 37 °C with a humidified atmosphere of 5% CO2 for 2 h and subsequently resuspended in FBS-free RPMI 1640 medium. A suspension containing 1 x 10^6^ cells in 100 µL was then loaded into the upper chamber of Transwell inserts (5 µm, Corning, Kaiserslautern, Germany). In the lower chambers, varying concentrations of CCL2 (e.g., 50 ng/mL, 100 ng/mL, 200 ng/mL, 400 ng/mL) in a total volume of 600 µL were added. The filters were subsequently transferred into the wells of the lower chamber. The 24-well plate containing the inserts was then placed in a 37°C incubator and 5% CO2 for 4 h. Cells that migrated into the lower chamber were carefully collected, washed once with PBS containing 0.5% BSA, and stained with Alexa Fluor 488-conjugated anti-FLAG antibody (DYKDDDDK tag Alexa-Fluor, company, # 701629RP488). The stained cells were then incubated on ice for 1 h at 4°C. After incubation, the cells were thoroughly washed with the same buffer and counted by flow cytometry (BD FACSymphony A1^TM^, BD Biosciences) using CountBright™ Absolute counting beads (Molecular Probes-Invitrogen, Karlsruhe, Germany).

*Cyclic adenosine monophosphate assay*

Human HEK293T cells were lifted with non-enzymatic cell stripper and resuspended in assay buffer at desired concentrations. Cyclic AMP (cAMP) assays were performed according to the manufacturer’s protocol using the MULTISCREENTM TR-FRET cAMP 1.0 No Wash Assay Kit (Multispan, Inc., Cat# MSCM01). The cells were treated with the cognate ligand of CCR2, CCL2, reconstituted to 30 μM concentration in sterile 0.1% BSA PBS, followed by addition of forskolin and incubation at 37 ^o^C for 20 min. The reaction was terminated by sequentially adding MULTISCREEN^TM^ Eu-labeled cAMP and MULTISCREEN^TM^ 650-labeled anti-cAMP antibody in lysis buffer. The plate was then incubated at room temperature for 30 min before reading fluorescent emissions at 620 nm and 665 nm with excitation at 314 nm on FlexStation III (Molecular Devices). The HEK293T cells stably expressing CCR2, HEK293T parental cells, and transiently transfected mutant CCR2 pools were used in the assay. Cells were stained with an anti-FLAG antibody at 2 μg/mL at 4^o^C for 1 h in the dark, followed by 3 washes in PBS plus 0.1% BSA and 0.2% sodium azide before being analyzed on a flow cytometer for surface receptor expression. 3,000 events were collected for each sample and the data were analyzed using CellQuest Pro (Becton Dickinson). The cAMP assay results are presented as “Ratio 665/620 x 10,000” (ratio of fluorescence at 665 nm and 620 nm x 10,000).

**Replication biobanks**

*TOPMed*

The Trans-Omics for Precision Medicine (TOPMed) program sponsored by the National Heart, Lung and Blood Institute (NHLBI) has generated deep whole genome sequencing (WGS) data in over 80 participating cohorts and over 150,000 participants. TOPMed whole geneome sequencing methods have been described fully previously^2^. Briefly, WGS was performed at the designated sequencing center for each included study. The sequencing data was aligned to human genome build GRCh38, and basic quality control measures were performed by the TOPMed Informatics Research Center. The data were subsequently released in different freezes. This analysis used Freeze 10. A total of 184,878 TOPMed, CCDG and 1000G samples underwent ≈30× WGS using DNA extracted from blood samples at designated sequencing centers for variant discovery. Basic quality control procedures were implemented by the TOPMed Informatics Research Center.

Of the 80 participating cohorts, 12 cohorts (**Supplemental Table S7**) have data on myocardial infarction and were made available for this study (n= 6,425 cases and 45,307 controls). TOPMed phenotype data was harmonized by the TOPMed Atherosclerosis Working Group to create prevalent coronary artery disease cases and control for each cohort. Cases were defined as individuals who experienced acute myocardial infarction according to hospital records or self-report. Controls were defined as non-cases who did not have documented angina, coronary artery revascularization and did not have possible coronary artery disease related death. One cohort, BioMe, had a high prevalence of peripheral arterial disease (PAD). Individuals with PAD were also removed from being considered a control. TOPMed WGS and phenotype data are available on dbGAP (study access numbers in Supplemental Table 1)).

The analysis was performed on Encore, a web-based tool to conducted large scale association testing with TOPMed sequencing data, using a Saige Logistic Mixed Model accounting for genetic relatedness through a kinship matrix (<https://doi.org/10.1038/s41588-018-0184-y>). The model adjusted for the following covariates: sex, self-identified race/ethnicity, TOPMed cohort, sequencing center, and ancestral principal components (PCs) 1 through 10. PCs were computed using PLINK (<http://pngu.mgh.harvard.edu/purcell/plink/>).

*DeCODE study*

Icelandic myocardial infarction cases were identified from a registry of individuals diagnosed at Landspitali University Hospital in Reykjavik, the only tertiary referral center in Iceland, during the years 1981 to 2022.  The criteria for Myocardial infarction diagnosis were defined as previously described in Helgadottir et al, 2007^3^ and genotyping and statistical analysis as described in Aegisdottir et al. 2023.^4^

*Penn Medicine Biobank*

The Penn Medicine Biobank (PMBB) is an academic healthcare system-based genomic and precision medicine cohort that links participant blood and tissue samples with associated health information. Procedures for recruitment, consent, data collection and genotyping are detailed elsewhere.^5^ Individuals of European ancestry with available whole-exome sequencing data were included in the analysis (n=43,723). Among whole exome variants in the CCR2 gene, we identified a total of 14 predicted LOF or potentially deleterious (REVEL > 0.5) variants, distributed across 68 heterozygous carriers and no homozygous carriers. Association testing against outcomes of interest was performed using Firth’s penalized logistic regression - adjusting for age, sex, and the first 5 principal components of ancestry. Myocardial infarction was defined as previously described (Pan-UKB team: <https://pan.ukbb.broadinstitute.org> 2020).

*Geisinger DiscovEHR-MyCode cohort*

The Geisinger Institutional Review Board approved this study (IRB No. 2019-0740) to meet “Non-human subject research” using de-identified information. All research was performed in accordance with relevant guidelines/regulations. The DiscovEHR(Geisinger) cohort is an integrated healthcare system located in central and northeastern Pennsylvania. Geisinger built and performs regular updates to the de-identified structured EHR database for research linked to the MyCode Community Health Initiative biorepository (Dewey FE et al). This research biobank is open to all Geisinger patients regardless of age or underlying disease. Informed consent was obtained from all subjects and/or their legal guardian(s) for the MyCode patients.

DNA Samples from 170,503 MyCode participants have been exome sequenced by IDT exome capture/Illumina HiSeq and genotyped using Infinium OmniExpress Exome array (Illumina), and GSA-24v1-0 array (Illumina) by the Regeneron Genetics Center(citation). For the exome sequencing data (WeCall/GLnexus), the patient-level quality metrics from the segment pVCF for *CCR2* gene were extracted. GQ(Genotype Quality) ≥ 30, DP(Depth of Coverage) ≥ 15, AD(Allelic Depth) ≥ 20, and SBPV(Strand Bias P-value) ≥ 10% were set as variant quality filters (Haggerty CM, et al). For array-based genotyping data, SNPs with minor allele frequency (MAF) ≤ 1%, significant deviation (p ≤ 1x10^-15^) from Hardy-Weinberg Equilibrium (HWE), and site-level missingness ≥ 1%, were removed. Array-based genotypes were imputed to the TOPMED reference panel (97,256 deeply sequenced genomes) with a GRCh38 build using the TOPMED Imputation Server (https://imputation.biodatacatalyst.nhlbi.nih.gov), which employed Eagle v2.4 and Minimac4 as the phasing and imputation algorithm, respectively. When completed, the imputed data in VCF files were retrieved from the server and merged sample-wise using bcftools (https://samtools.github.io/bcftools/bcftools.html) in 5MB genomic regions. Only the imputed variants with the info score ≥0.7 were considered for the following analysis. A set of LD-pruned (200 variant windows, 50 variant sliding windows and r^2^ < 0.25) common variants (~1M) were applied for PCA (PLINK2.0) and 5 major PCs were included as covariates for the association study.

Geisinger billing codes for diagnostics at any encounter setting were mapped to the *International Classification of Disease (ICD)*-related ICD9 and ICD10 codes. Demographic information was extracted from the structured EHR. ICD-9 and ICD-10 diagnostic codes were combined and mapped to 1,866 hierarchical PheCodes, each representing a specific disease phenotype. Study participants were labeled with a PheCode if they had two or more of the PheCode-specific ICD codes. “Case” was defined as all study participants with the PheCode of interest, and “control” was defined as all study participants without the PheCode of interest. Sex information was included during the mapping process so that sex-specific PheCodes would not be mislabeled.

Only the individuals (n = 167,720) from the MyCode sample with at least two encounters for more than 3 months apart in the Geisinger EHR were recruited for the following association study. 145,832 participants had genetically determined EUR ancestry. The index age was calculated by the subtraction of the last active date (primarily last encounter date) and the birth date. The sex, index age, and five major principal components (PCs) were included as covariates in the logistic regression model with Firth’s correction (R logistf package). Other risk factors were extracted from the Geisinger EHR without missing value.

*Million Veterans Programme*

GWAS was performed for each trait within ancestry group using mixed models implemented in SAIGE and adjusting for age, sex, and the first 10 PCs. The examined variant was directly genotyped. Further information about the genotyping approach is provided elsewhere.^6^

*Mass General Brigham Biobank*

Data in the MGB biobank consisted of ~36000 individuals, contained within 6 batches within 8 separate files. The reference genome used for the genotyping was GRCh37. Quality control was initially performed on individual datasets as follows: Deletion of SNPs/individuals with missingness >10%, deletion of sex discrepancies, deletion of variants where Hardy-Weinberg equilibrium < 10^-15 or MAF < 1%, deletion of individuals where heterozygosity rate >3 sd from mean, deletion of subjects that are related at pi-hat >0.2. After merging of datasets, a second quality control was performed with the same criteria except no deletion of individuals with missingness and thresholds of HWE was set at 10^-6. After pruning of variants with high LD, a dataset consisting of 31098 individuals with +/- 86K variants was used for calculation of the first 5 ancestral principal components. Myocardial infarction (MI) and coronary artery disease (CAD) were assessed using the MGB biobank query tool, where both were defined using ICD-9 and 10 codes. Below the effect which corresponds to the odds ratio derived from logistic regression adjusted for age, sex, batch number and the first 5 ancestral principal components.

**SUPPLEMENTARY FIGURES**

**Supplementary Figure S1. Migration assay with monocyte-lowering *CCR2* variants on CCL2-elicited monocyte chemotaxis.** Impact of transfection of *CCR2*-knockout human monocytes (THP-1 cell line) with M249K and W165S *CCR2* mutants (lower row), as compared to no transfection or transfection with wild-type *CCR2* (upper row) on chemotaxis to CCL2 in a migration assay. info on stats missing – P value/** etc.; ANOVA xxx (“one-way ANOVA?! Posthoc multiple comparisons??, ns: p>0.05, *** p<0.001, ****p<0.0001

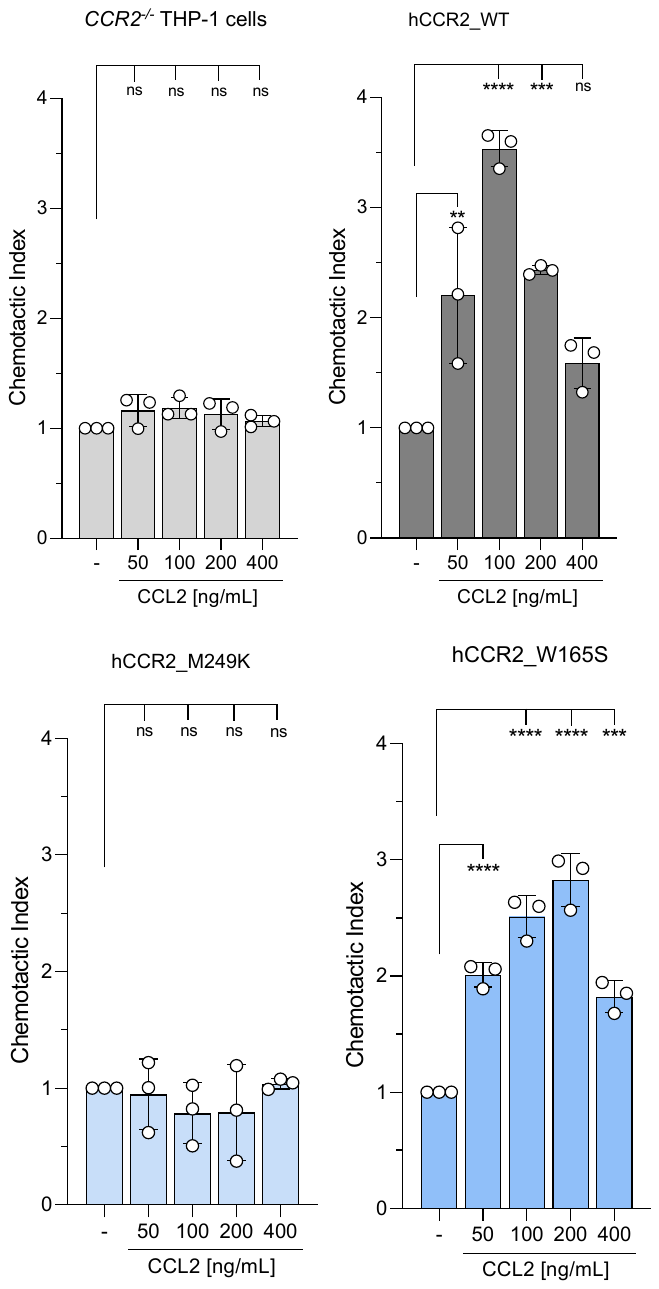

**Supplementary Figure S2.** Meta-analysis of six external cohorts for the effect of the M249K variant (767 carriers) on risk of (A) myocardial infarction (N=1,062,595, 91,420 cases) and (B) coronary artery disease (N= 956,990, 160,977 cases). All cohorts are adjusted for age, sex, and the first 5 ancestral principal components.

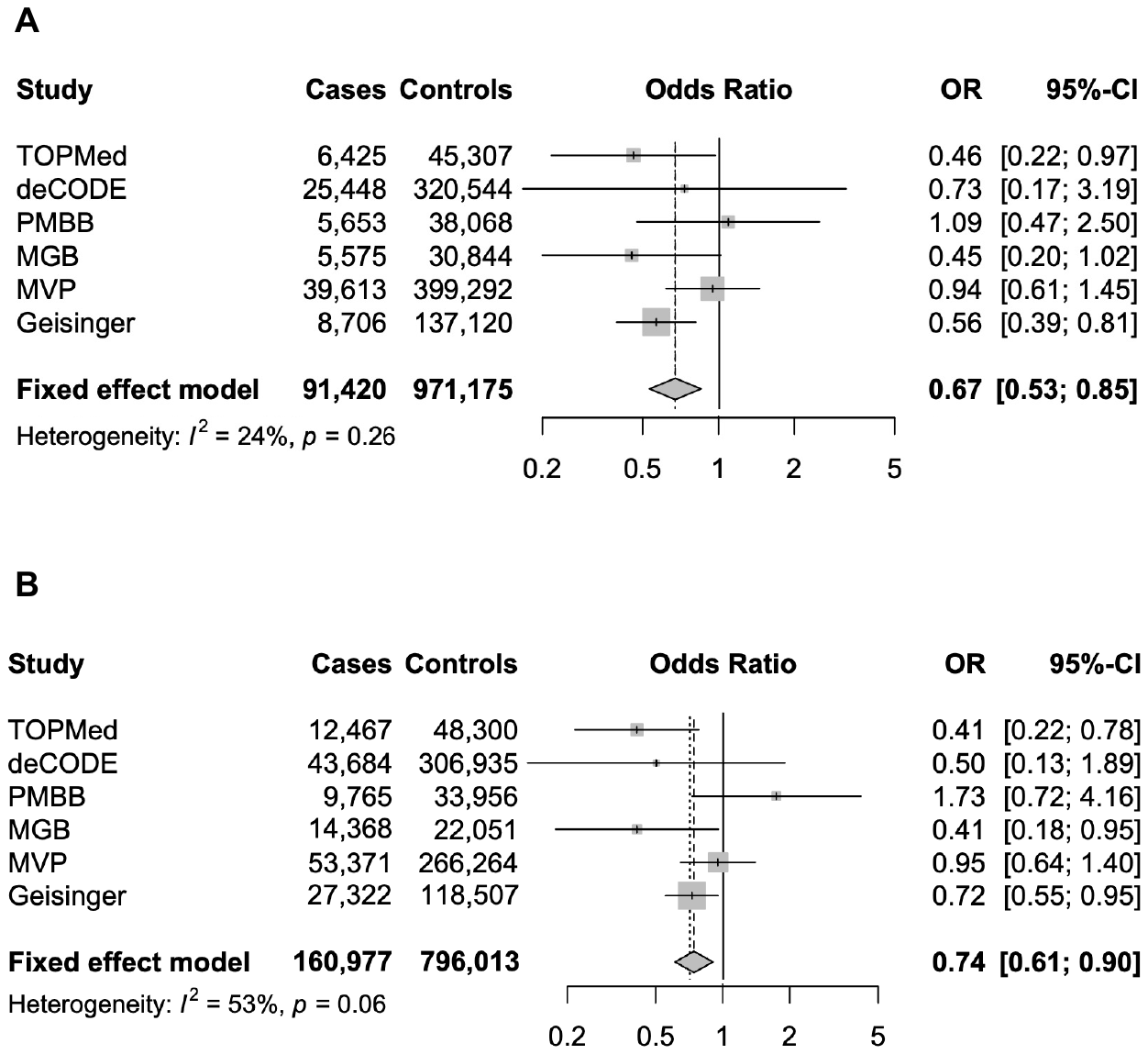
